# Impact of long-COVID on health-related quality of life in Japanese COVID-19 patients

**DOI:** 10.1101/2021.09.27.21264225

**Authors:** Shinya Tsuzuki, Yusuke Miyazato, Mari Terada, Shinichiro Morioka, Norio Ohmagari, Philippe Beutels

**Author notes:** Corresponding author: Shinya Tsuzuki, MD, MSc, National Center for Global Health and Medicine, 1-21-1 Toyama, Shinjuku-ku, Tokyo 162-8655, Japan, University of Antwerp, Prinsstraat 13, 2000 Antwerpen, Belgium.

## Abstract

**Background:** The empirical basis for a quantitative assessment of the disease burden imposed by long-COVID is currently scant. We aimed to assess the disease burden caused by long-COVID in Japan.

**Methods:** We conducted a cross sectional self-report questionnaire survey. The questionnaire was mailed to 530 eligible patients, who were recovered from acute COVID-19 in April 2021. Answers were classified into two groups; participants who have no symptom and those who have any ongoing symptoms that lasted longer than four weeks at the time of the survey. We compared health-related quality of life scores estimated by the EQ-5D-3L questionnaire between these two groups after adjusting basic characteristics of the participants by propensity score matching.

**Results:** 349 participants reported no symptoms and 108 reported any symptoms at the time of the survey. The participants who reported any symptoms showed a lower value on a Visual Analogue Scale (median 70 [IQR 60-80]) and on the EQ-5D-3L (median 0.81 [IQR 0.77-1.0]) than those reporting no symptoms (median 85 [IQR 75-90] and 1.0 [IQR 1.0-1.0], respectively). After adjusting for background characteristics, these trends did not change substantially (Visual Analog Scale: median 70 [IQR 60-80] vs 80 [IQR 77-90], EQ-5D-3L: median 0.81 [IQR 0.76-1.0] vs 1.0 [IQR 1.0-1.0]).

**Conclusions:** Due to their long duration, long-COVID symptoms represent a substantial disease burden expressed in impact on health-related quality of life.

**Trial registration:** Not applicable.

## Background

Coronavirus disease 2019 (COVID-19) caused by the SARS-CoV-2 virus, has become a global health threat [1]. Not only its acute phase of disease, but so-called “long-COVID” is also a cause of substantial disease burden [2,3]. A systematic review reported that 80% of patients developed one or more long-term symptoms and the prevalence of 55 long-term effects of COVID-19 [4].

There is no clear definition of long-COVID so far, however, the National Institute for Health and Care Excellence (NICE) in The UK defined it as “signs and symptoms that develop during or following an infection consistent with covid-19 and which continue for more than four weeks and are not explained by an alternative diagnosis” [5]. This term includes ongoing symptomatic COVID-19, from four to 12 weeks post-infection, and post-COVID-19 syndrome, beyond 12 weeks post-infection [6].

The symptoms of long-COVID are various and often different from the acute phase of COVID-19. Miyazato and colleagues reported that the mean time from COVID-19 symptom onset to the emergence of alopecia was 58.6 days and one of patients presented dysosmia after 92 days after symptom onset [7]. Other symptoms such as general fatigue [8,9], respiratory symptoms [10,11], cognitive and mental health disorder [12,13], and so forth [14,15] have been reported as long-COVID.

Considering its chronic phase, the disease burden of COVID-19 should be larger than that of other respiratory infections due to length and variety of the symptoms. However, the empirical basis for a quantitative assessment of the disease burden imposed by long-COVID is currently scant. An important element towards disease burden assessment is health-related quality of life impact. The objective of the present study is to collect and analyse empirical information on the health-related quality of life (HRQoL) due to long-COVID.

## Methods

### Settings

We conducted a survey in which a self-report questionnaire was mailed to eligible patients who had recovered from COVID-19 in April 2021 with two reminders 2 weeks and 1 month later. Potential participants were recruited from patients who had recovered from COVID-19 and visited the outpatient service of the Disease Control and Prevention Center (DCC) in the NCGM in order to obtain pre-donation screening test for COVID-19 convalescent plasmapheresis (Another study named “Collection and antibody measurement of Convalescent plasma foreseeing the use for COVID-19 treatment”). 530 patients who had recovered from COVID-19 (70 out of 448 needs supplementary oxygen support) and visited the outpatient service of the Disease Control and Prevention Center (DCC) in National Center for Global Health and Medicine (NCGM), Tokyo, Japan from 1^st^ February 2020 to 31^st^ March 2021. Participation in the survey was voluntary and not anonymous. Participants were requested to complete and return the questionnaire. 457 of 530 (86.2%) patients completely answered the questionnaire and were included in the analysis.

### Statistical analysis

Answers were classified into two groups; participants who have no symptom and those who have any ongoing symptoms that lasted longer than four weeks at the time of the survey. We compared visual analogue scale (VAS) and HRQoL values estimated by the EQ-5D-3L questionnaire [16] by Japanese value set [17] between two groups using Mann-Whitney U test, after adjusting basic characteristics of the participants by one-to-one propensity score matching (nearest neighbour pair matching, caliper = 0.2) calculated by multivariate logistic regression model predicting the likelihood of having ongoing symptoms [18,19]. We included age, sex, BMI, smoking, drinking, hypertension, diabetes, chronic obstructive lung diseases, malignancy, use of antivirals, use of systemic steroids, and severe COVID-19 disease during admission (use of mechanical ventilation or extracorporeal membrane oxygenation during admission, according to the definition by a report of national registry data in Japan [20]). The standardized mean difference was used to measure covariate balance, and an absolute standardized difference above 20% was interpreted as a meaningful imbalance. Two-sided p values of <0.05 were considered to show statistical significance. All analyses were conducted by R, version 4.0.5 [21].

### Ethics approval

According to local ethical guidelines, responses to questionnaire were regarded as patient consent. This study was reviewed and approved by the Ethics Committee of the Center Hospital of the NCGM (NCGM-G-004121-00).

## Results

Table 1 shows the basic characteristics of the participants. 457 patients recovered from acute phase of COVID-19 and 108 of them presented at least one symptom. The proportion of female was larger in “Any symptom” group than that in “No symptom” group. There was no substantial difference between the two groups in terms of their age and medical history. Crude comparison of VAS and QOL showed that “Any symptom” group had lower VAS and QOL values than the “No symptom” group did (VAS: 70 vs 85, QOL: 0.81 vs 1.0, respectively).

**Table 1.**
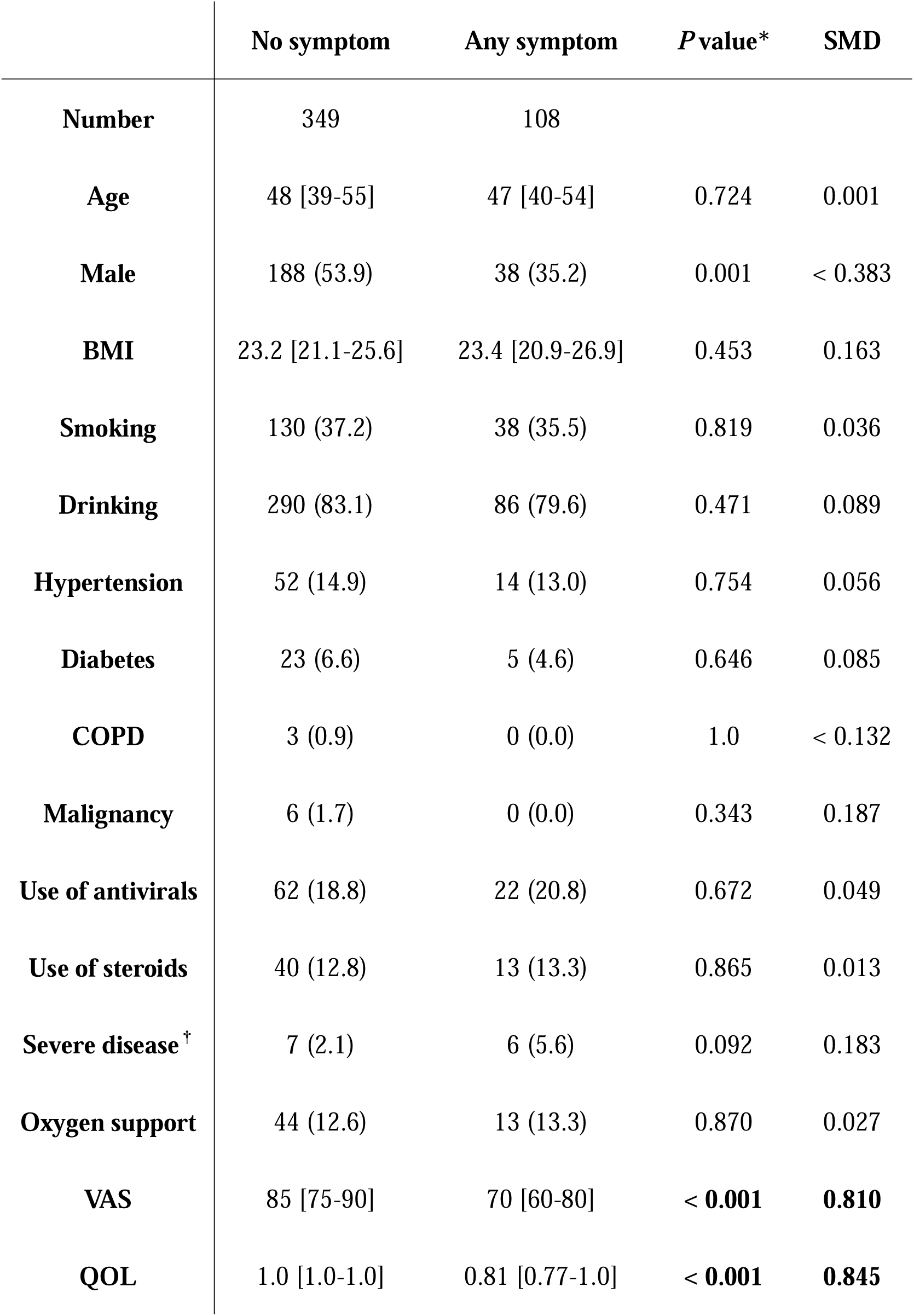
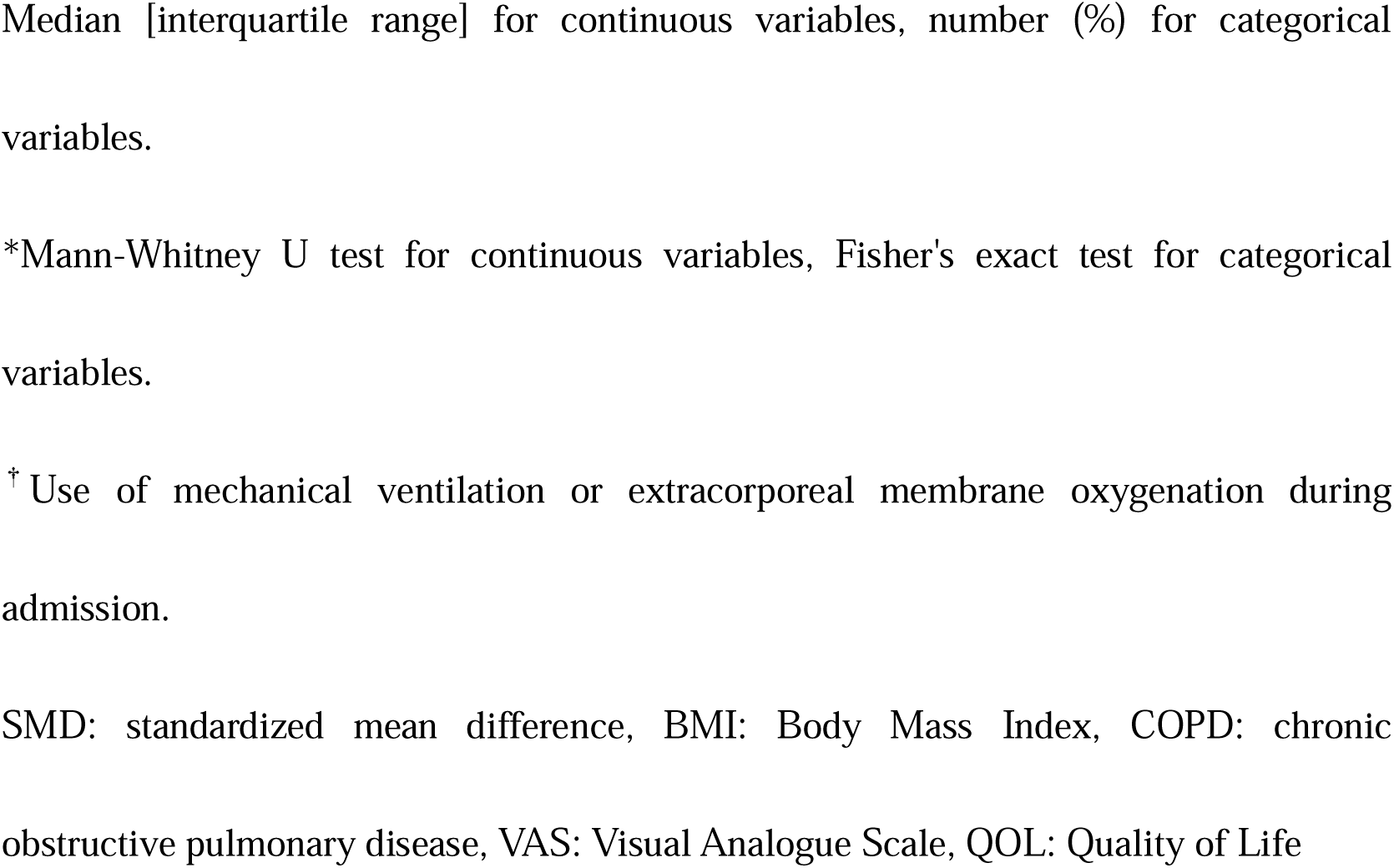
Characteristics of participants.

|  | No symptom | Any symptom | *P* value* | SMD |
| --- | --- | --- | --- | --- |
| **Number** | 349 | 108 |  |  |
| **Age** | 48 [39-55] | 47 [40-54] | 0.724 | 0.001 |
| **Male** | 188 (53.9) | 38 (35.2) | 0.001 | < 0.383 |
| **BMI** | 23.2 [21.1-25.6] | 23.4 [20.9-26.9] | 0.453 | 0.163 |
| **Smoking** | 130 (37.2) | 38 (35.5) | 0.819 | 0.036 |
| **Drinking** | 290 (83.1) | 86 (79.6) | 0.471 | 0.089 |
| **Hypertension** | 52 (14.9) | 14 (13.0) | 0.754 | 0.056 |
| **Diabetes** | 23 (6.6) | 5 (4.6) | 0.646 | 0.085 |
| **COPD** | 3 (0.9) | 0 (0.0) | 1.0 | < 0.132 |
| **Malignancy** | 6 (1.7) | 0 (0.0) | 0.343 | 0.187 |
| **Use of antivirals** | 62 (18.8) | 22 (20.8) | 0.672 | 0.049 |
| **Use of steroids** | 40 (12.8) | 13 (13.3) | 0.865 | 0.013 |
| **Severe disease<sup>†</sup>** | 7 (2.1) | 6 (5.6) | 0.092 | 0.183 |
| **Oxygen support** | 44 (12.6) | 13 (13.3) | 0.870 | 0.027 |
| **VAS** | 85 [75-90] | 70 [60-80] | **< 0.001** | **0.810** |
| **QOL** | 1.0 [1.0-1.0] | 0.81 [0.77-1.0] | **< 0.001** | **0.845** |

Median [interquartile range] for continuous variables, number (%) for categorical variables.
\*Mann-Whitney U test for continuous variables, Fisher's exact test for categorical variables.
<sup>†</sup> Use of mechanical ventilation or extracorporeal membrane oxygenation during admission.
SMD: standardized mean difference, BMI: Body Mass Index, COPD: chronic obstructive pulmonary disease, VAS: Visual Analogue Scale, QOL: Quality of Life

Table 2 describes the characteristics of the data after propensity score matching. 91 pairs included in the matched data and 17 of “Any symptom” group were discarded. Adjusted comparison of VAS and QOL showed a similar trend to the results of crude comparison. Both VAS and QOL were lower in the “Any symptom” group (VAS: 70 vs 80, QOL: 0.81 vs 1.0, respectively).

**Table 2.**
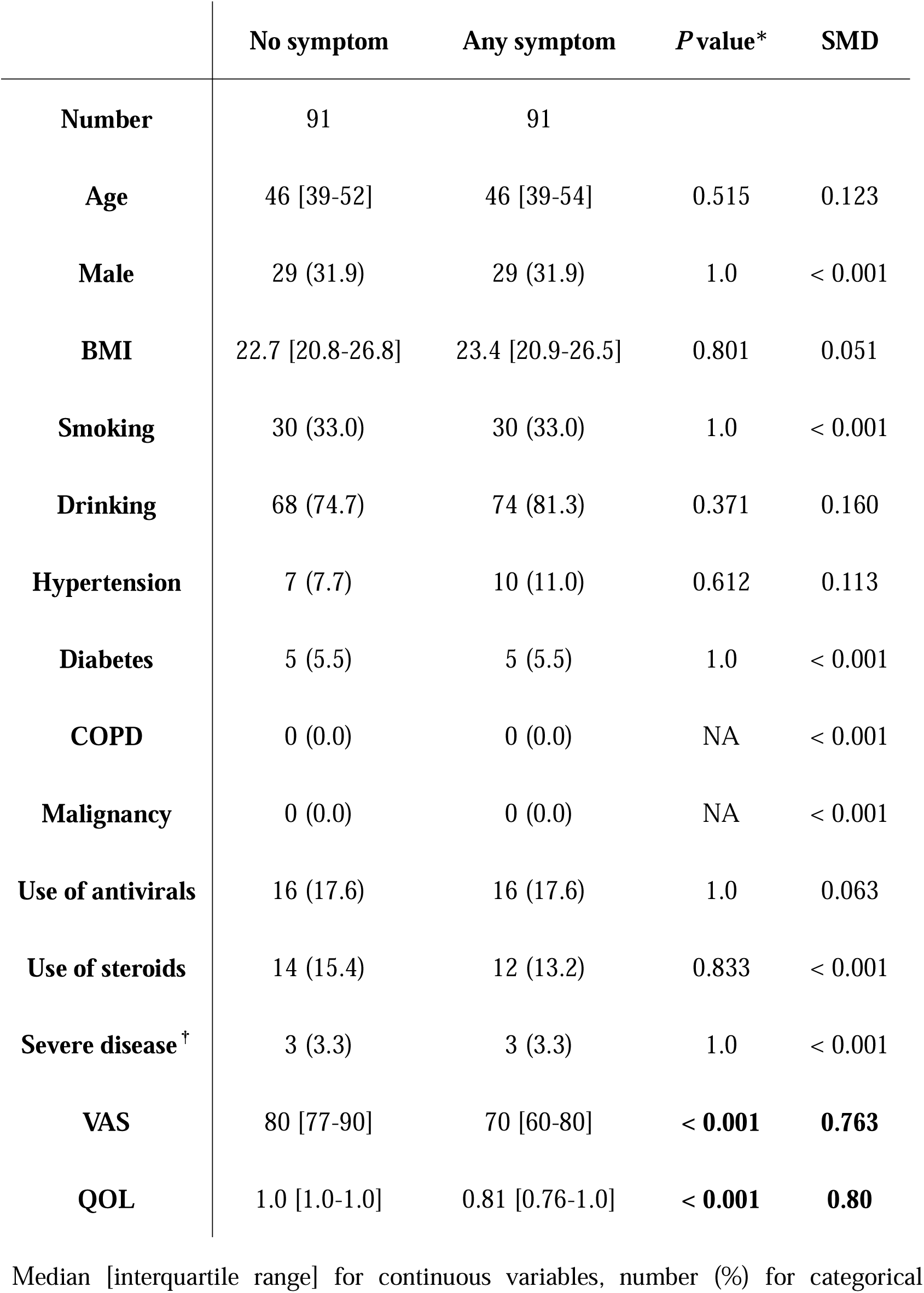
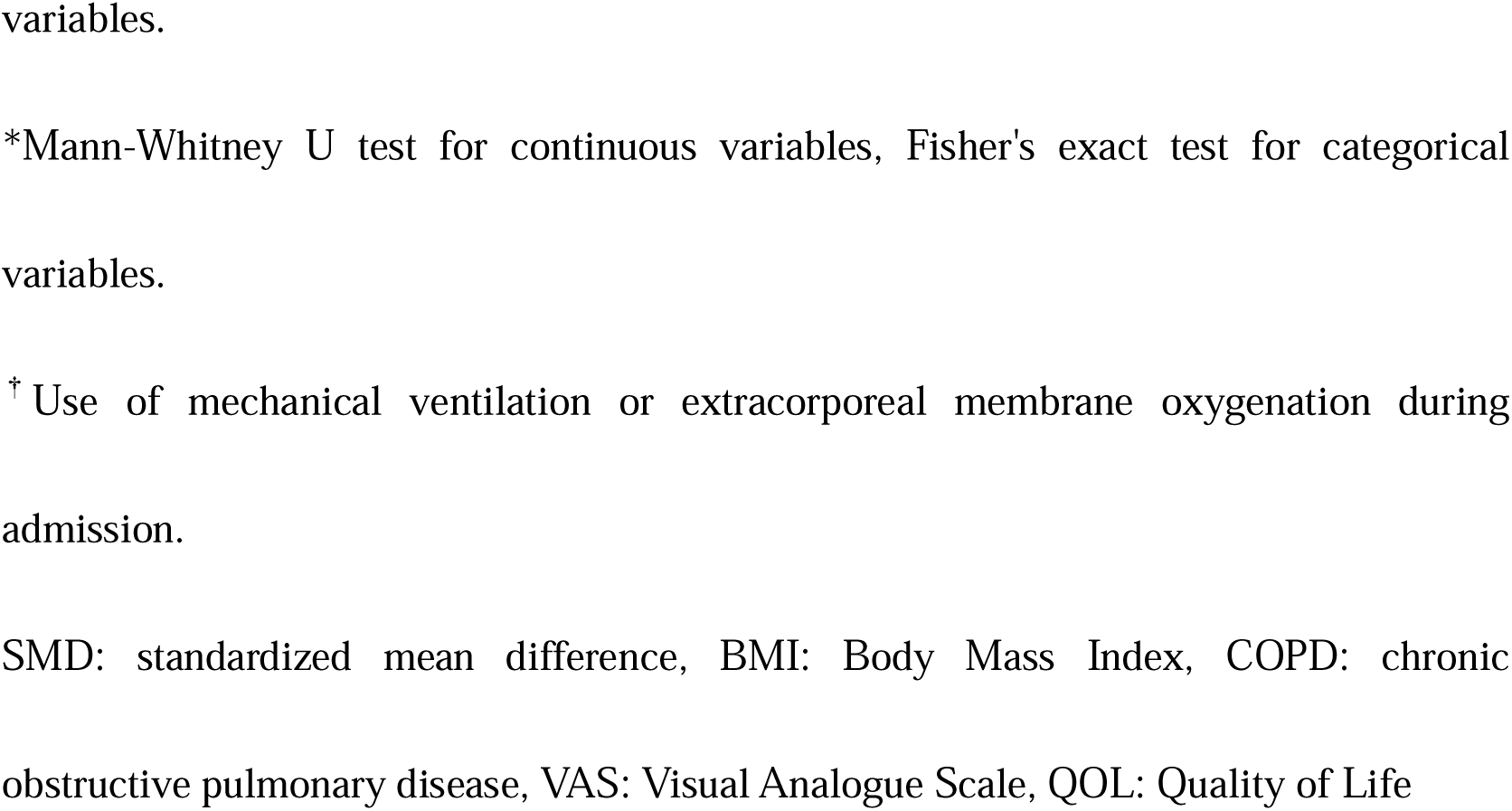
Characteristics of participants after propensity score matching.

|  | No symptom | Any symptom | *P* value* | SMD |
| --- | --- | --- | --- | --- |
| **Number** | 91 | 91 |  |  |
| **Age** | 46 [39-52] | 46 [39-54] | 0.515 | 0.123 |
| **Male** | 29 (31.9) | 29 (31.9) | 1.0 | < 0.001 |
| **BMI** | 22.7 [20.8-26.8] | 23.4 [20.9-26.5] | 0.801 | 0.051 |
| **Smoking** | 30 (33.0) | 30 (33.0) | 1.0 | < 0.001 |
| **Drinking** | 68 (74.7) | 74 (81.3) | 0.371 | 0.160 |
| **Hypertension** | 7 (7.7) | 10 (11.0) | 0.612 | 0.113 |
| **Diabetes** | 5 (5.5) | 5 (5.5) | 1.0 | < 0.001 |
| **COPD** | 0 (0.0) | 0 (0.0) | NA | < 0.001 |
| **Malignancy** | 0 (0.0) | 0 (0.0) | NA | < 0.001 |
| **Use of antivirals** | 16 (17.6) | 16 (17.6) | 1.0 | 0.063 |
| **Use of steroids** | 14 (15.4) | 12 (13.2) | 0.833 | < 0.001 |
| **Severe disease<sup>†</sup>** | 3 (3.3) | 3 (3.3) | 1.0 | < 0.001 |
| **VAS** | 80 [77-90] | 70 [60-80] | **< 0.001** | **0.763** |
| **QOL** | 1.0 [1.0-1.0] | 0.81 [0.76-1.0] | **< 0.001** | **0.80** |

Median [interquartile range] for continuous variables, number (%) for categorical
variables.
\*Mann-Whitney U test for continuous variables, Fisher's exact test for categorical variables.
<sup>†</sup> Use of mechanical ventilation or extracorporeal membrane oxygenation during admission.
SMD: standardized mean difference, BMI: Body Mass Index, COPD: chronic obstructive pulmonary disease, VAS: Visual Analogue Scale, QOL: Quality of Life

Table 3 describes the characteristics of “long-COVID” symptoms. We defined “long-COVID” as the status in which any symptoms attributed to SARS-nCoV-2 infection lasting more than four weeks in the present study. 201 of 457 (44.0%) participants reported at least one symptom after four weeks have passed since their symptom onset due to COVID-19. The most common symptom of long-COVID was general fatigue. 58 of 457 (12.7%) participants have had general fatigue longer than four weeks. The second most common symptom was alopecia. 55 of 457 (12.0%) participants have experienced hair loss worse than usual.

**Table 3.** Details of symptoms lasted longer than four weeks in the participants.

|  | **Number** | **Duration (days)** |
| --- | --- | --- |
| **Number of symptoms** | 457 |  |
| **At least 1** | 201 (44.0) |  |
| **1** | 73 (16.0) |  |
| **2** | 46 (10.1) |  |
| **3** | 47 (10.3) |  |
| **4 and more** | 34 (7.4) |  |
| **Fatigue** | 58 (12.7) | 50 [30-60] |
| **Hair loss** | 55 (12.0) | 60 [30-90] |
| **Cough** | 54 (11.8) | 40 [30-60] |
| **Dysosmia** | 47 (10.3) | 45 [30-60] |
| **Dysgeusia** | 47 (10.3) | 35 [30-60] |
| **Shortness of breath** | 36 (7.9) | 42.5 [30-60] |
| **Loss of concentration** | 34 (7.4) | 40 [30-90] |
| **Depression** | 29 (6.3) | 40 [30-60] |

Absolute number (%) for the number of participants, median [interquartile range] for the duration of symptoms.

Figure 1 and 2 show a violin plot of VAS and QOL value, respectively. The “Any symptom” group showed greater variance in both indicators.

**Figure 1.**
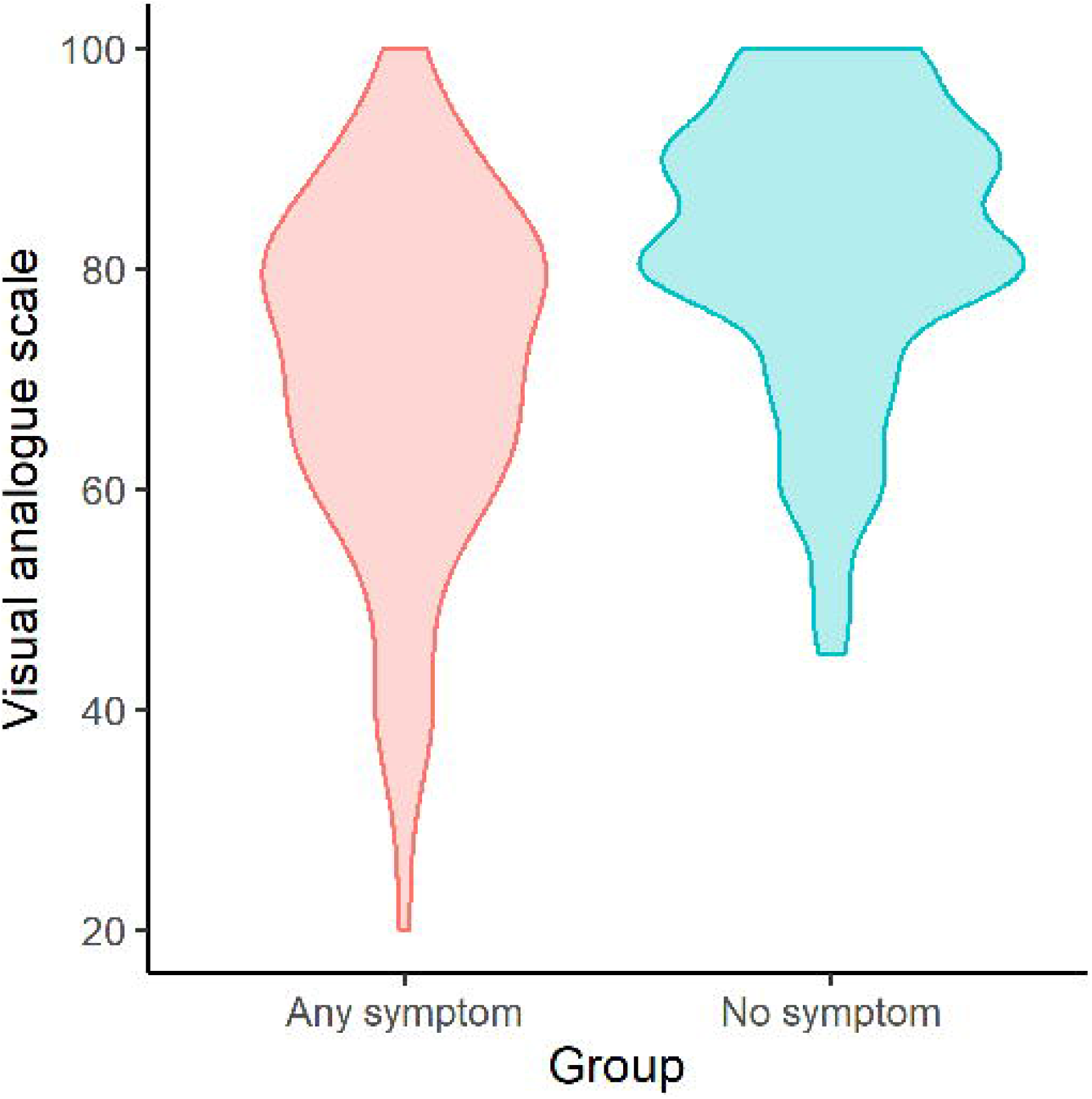
Violin plot of visual analogue scale. Red colour represents “No symptom” group and blue colour represents “With symptom” group.

**Figure 2.**
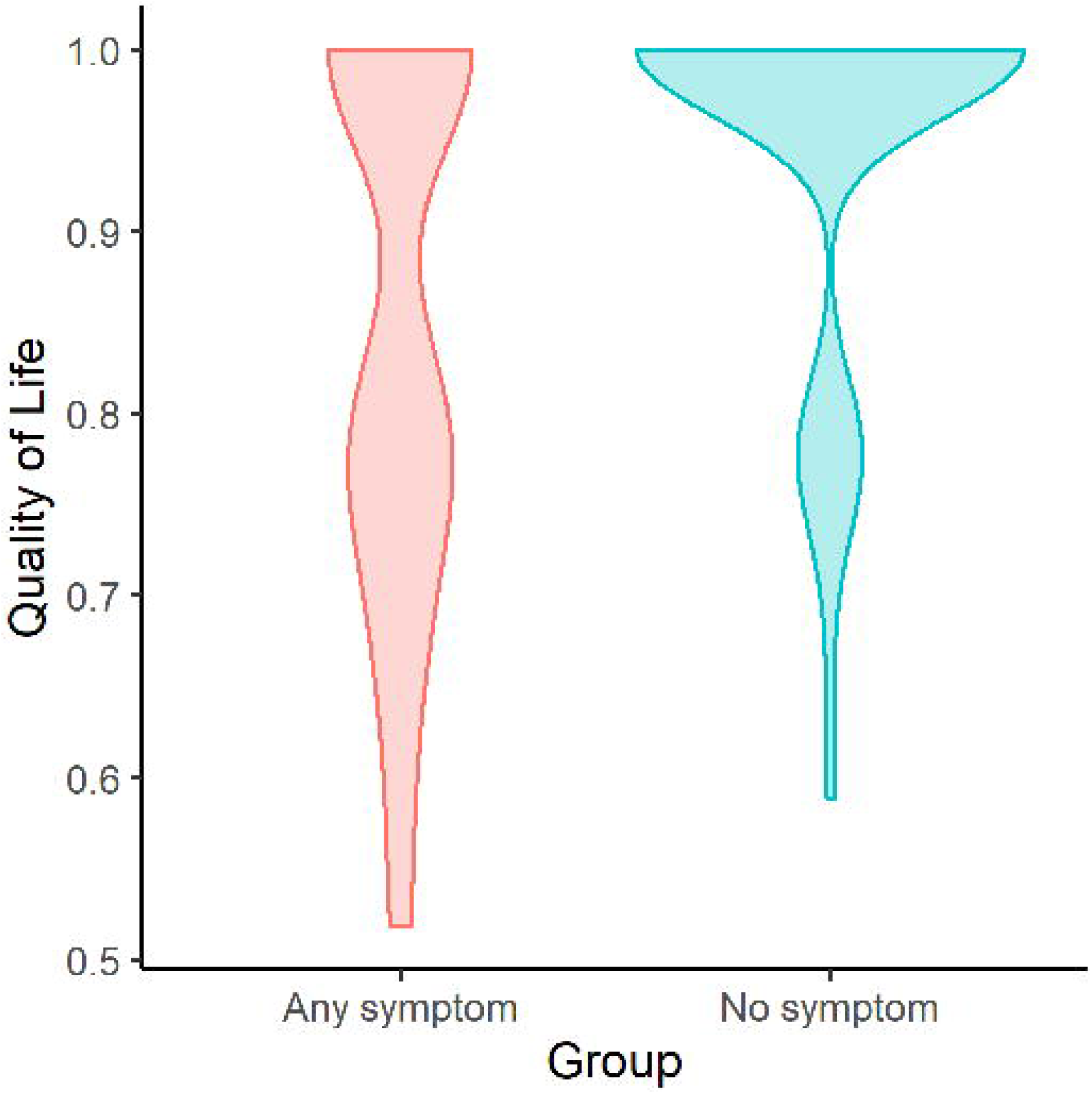
Violin plot of Quality of Life. Red colour represents “No symptom” group and blue colour represents “With symptom” group.

## Discussion

The present study demonstrated that the phenomenon we called “long-COVID” can impair the HRQOL substantially. This would be another important aspect of COVID-19 to consider because it implies a heavier disease burden to us than other influenza like illnesses (ILIs) do, not only due to its severity but also the characteristics of its chronic phase. In the first place, COVID-19 showed higher case-fatality than other ILIs [22–24]. Additionally, it might cause a substantial burden through accumulated mild disease only.

Furthermore, the frequency and the duration of symptoms due to “long-COVID” are also noteworthy. Our results showed that nearly half of the patients who recovered from acute COVID-19 (201/457) experienced any symptoms lasting more than four weeks. As for patients who required supplementary oxygen support, 32 out of 70 (45.7%) presented any symptoms longer than four weeks. The precise duration of such symptoms was not obvious because more than 100 participants reported that their symptoms were still ongoing, nevertheless, we can say that the symptoms attributed to “long-COVID” often continue several months. Albeit the HRQOL value of the participants who have any “long-COVID” symptoms was higher than that of the acute phase of other ILIs reported by a previous study in Japan (0.81 vs 0.66, respectively) [25], QOL lost attributed to “long-COVID” should be greater than that due to the acute phase of other ILIs because of its duration.

These results suggest that prevention is more important in COVID-19 countermeasures than other ILIs because effective treatment of “long-COVID” is not clearly established yet [6,26]. Although there is no doubt that vaccination against SARS-CoV-2 will reduce the risk of fatal and severe COVID-19 [27–29], its effectiveness against “long-COVID” is not demonstrated yet. This may provide an additional incentive to prevent SARS-CoV-2 infection even in the absence of known risk factors of severe illness.

There are several limitations in our study. First, our results are based on the questionnaire survey then there are some recall biases in participants’ responses. Similarly, the potential participants were enrolled from the visitors of outpatient department at the national center hospital of infectious diseases in Japan, then the study population might be influenced by selection biases. In addition, we could not take “new variants” into consideration. The difference in severity, infectiousness, and so forth between such new variants and old ones were already reported [30–32], however, there is no solid evidence about the frequency and the severity of “long-COVID” symptoms in new variants. This should be the subject of future study.

In addition, we should be careful about the representativeness of the data when we interpret the results because our survey includes a comparatively small number of participants from Japan. However, the response rate of our survey was extremely high (86.2%), and non-response bias may therefore be limited. Furthermore, we compared VAS and EQ-5D-3L values after adjusting participants’ background by propensity score matching.

## Conclusions

What we call “long-COVID” brings us substantial disease burden in addition to the burden attributed to the acute phase of COVID-19. This additional burden makes the whole disease burden of COVID-19 heavier, making prevention strategies all the more important. The influence of vaccination and variants on “long-COVID” should be examined in the near future.

## Data Availability

The data that support the findings of this study are available upon request from the corresponding author. The data are not publicly available due to privacy or ethical restrictions.

## Declarations

### Ethics approval

This study was reviewed and approved by the Ethics Committee of the Center Hospital of the NCGM (NCGM-G-004121-00)

### Consent for publication

According to local ethical guidelines, responses to questionnaire were regarded as patient consent.

### Authors’ contributions

Shinya Tsuzuki: Conceptualization, Funding Acquisition, Data Curation, Formal Analysis, Methodology, Visualization, Original Draft Preparation. Yusuke Miyazato: Data Curation, Review and Editing. Mari Terada: Data Curation, Review and Editing. Shinichiro Morioka: Data Curation, Project Administration, Review and Editing. Norio Ohmagari: Conceptualization, Funding Acquisition, Project Administration, Review and Editing. Philippe Beutels: Supervision, Project Administration, Review & Editing.

## Acknowledgments

We thank all the people who participated in our survey and study, “Collection and antibody measurement of Convalescent plasma foreseeing the use for COVID-19 treatment”.

